# A Systematic Review and Meta-Analysis of the Incidence Rate of Takayasu Arteritis

**DOI:** 10.1101/2020.12.10.20246942

**Authors:** Megan Rutter, Jonathan Bowley, Peter C. Lanyon, Matthew J. Grainge, Fiona A. Pearce

## Abstract

**Objectives:** Takayasu arteritis (TAK), is a rare autoimmune rheumatic disease causing large vessel vasculitis. Onset is typically between the ages of 20-30. It is associated with substantial morbidity and mortality, notably due to its effects on the cardiovascular system. It has a poorly understood global epidemiology. Our objective was to systematically review the available evidence in order to calculate the incidence rate of TAK.

**Methods:** Three databases (Medline, PubMed and Embase) were searched in November 2019 and the results were screened by two reviewers. A random effects meta-analysis was then conducted in R to calculate the overall incidence rate. Heterogeneity was assessed using I^2^. The quality of the studies was assessed using an adapted Newcastle-Ottawa scale. Further sub-group analyses were performed by quality, sex, research setting and geographical location. Publication bias was assessed using a Begg’s funnel plot.

**Results:** The incidence rate for TAK per million person-years with 95% confidence intervals was 1.11 per million person years (95% CI 0.75 – 1.65). The heterogeneity in the data was extremely high in all analyses, which suggests that there was considerable variation in incidence rates across the different populations studied. TAK was found to be more common in women (incidence rate 2.01 per million person-years, 95% CI 1.39-2.90).

**Conclusions:** TAK is an extremely rare disease. It affects women more commonly than men. There is considerable variation in the incidence rate between populations. We suggest that future research should focus on discrete populations in order to better identify genetic and environmental risk factors.

## Introduction

Takayasu Arteritis (TAK) is a rare large-vessel vasculitis, which predominantly affects the aorta and its main branches. The disease and its sequelae lead to significant morbidity and mortality and may necessitate long-term immunosuppressive treatment (1, 2). The global epidemiology of TAK remains unknown. Prevalence seems to vary geographically and appears highest in Asia(3, 4).The first-line treatment for TAK is corticosteroids, with the addition of a disease modifying anti-rheumatic agent (DMARD) if there is a poor response to treatment(5). Tocilizumab, a monoclonal antibody against the IL-6 receptor, is now in use as a third line therapy in some countries including England (5). Many other drug pathways are under investigation, including TNF inhibition and JAK inhibition.

The lack of clarity on the basic epidemiology of TAK has many implications, including on the design of diagnostic pathways, creation of specialised services, funding of high-cost drugs and on clinical trial recruitment. We aimed to perform a systematic review and meta-analysis of the incidence of Takayasu arteritis world-wide.

## Methods

The method for this systematic review was guided by the Preferred Reporting Items for Systematic reviews and Meta-Analyses (PRISMA) statement for systematic reviews(6). The study protocol was registered on PROSPERO (an international prospective register of systematic reviews) on 9^th^ December 2019 with registration number CRD42019138795. Incidence rate for Takayasu arteritis was the primary outcome measure.

To meet the inclusion criteria, studies had to be cohort or retrospective cohort studies, report the incidence rate of TAK, and be published in English. Studies published prior to 1990, conference abstracts, reviews, and studies exclusively focusing on paediatric disease were excluded.

The academic databases used for the systematic review were MEDLINE (MEDLINE 1946 to present (OVID)), Embase (OVID) and PubMed. Key words relating to the population and outcome of interest were developed using synonyms for each disease and their respective medical subject headings (MeSH). A pilot search was done in Medline to ensure specificity of the search. A list of seminal papers was used to test the sensitivity of the search and all were found. Subsequent searches with key words were done with search terms developed for MEDLINE and further adapted for the other databases. Searches were carried out in November 2019. The search terms for Medline and PubMed were (Takayasu.mp.) AND (*incidence/ OR incidence.mp.) and for Embase (Takayasu.mp. OR aortic arch syndrome/) AND (incidence.mp. OR *incidence/).

Outputs from the literature search were screened in a three-stage logical manner, namely: title and abstract screening, full text screening and extraction. Two reviewers independently screened the retrieved studies. Conflicts in the study selection process were resolved by dialogue. In one case where the full text was not immediately available, the text was obtained following correspondence with the author.

The reviewers assessed the quality of selected studies using the Newcastle-Ottawa Scale, adapted to fit this review (Online Supplement, Appendix 1). The maximum number of points available was 5. Papers were grouped by number of points achieved, rather than in to high- or low-quality categories.

Data extraction was conducted in parallel by two reviewers (JB and FP). Data regarding study details (author, year of publication), study characteristics (design, setting) and participant data (population size, male:female ratio, mean age at diagnosis, incidence rate for diseases studied and the corresponding 95% confidence intervals) were extracted. The data extraction form used is shown in Appendix 2 (Online Supplement). Discrepancies were settled via dialogue.

The incidence rates were expressed per million person-years. Where papers didn’t provide the 95% confidence intervals, they were calculated using the formulae below, where D is the number of cases of the disease.

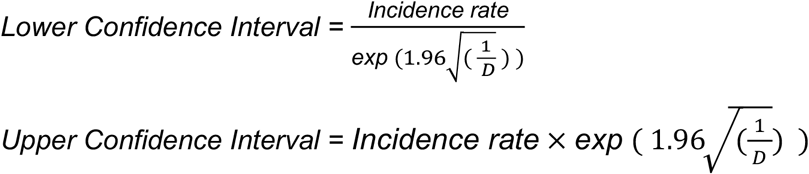

The distribution of incidence rates and 95% confidence intervals obtained from the studies were examined using a forest plot. Pooled estimates of the incidence rate were used to determine an overall rate using a random effects model. The log of the incidence rate and confidence intervals were calculated to allow for meta-analysis.

Assessment of heterogeneity was performed using the I^2^ statistic. I^2^ >50% was taken to represent moderate heterogeneity, and I^2^ >75% to represent high heterogeneity. Publication bias within studies was assessed with a funnel plot using a random effects model.

Subgroup analysis was employed to explore the effect of geographical region, sex, and the quality of the paper on incidence.

Many papers that reported the sex of the patients did not report the overall proportion of men and women in their populations. Where this was the case, a ratio of 50:50 was used as the estimated male:female ratio in the population. When calculating the effect of sex on the incidence, we found that several studies reported no cases among men. In this circumstance, in order to make meta-analysis possible, it is usual to put a rate slightly above zero in the group with no cases(7). Therefore, where the incidence rate in the male sub-group was zero, an incidence rate of 0.5 per million person/years was used in order to allow meta-analysis. We must also consider in the case of a rare outcome, whether arbitrarily adding to the numerator could, under random effects, cause an upward bias to the pooled estimate if we add 0.5 (or alternative quantity) to the zero event number for studies where a small denominator would mean that experiencing ≥1 events is unlikely. We took the pooled estimate for the annual incidence of 0.28/million in males and applied this to the male denominators for the studies by Romero-Gomez and Nesher to estimate the expected number of events and then applied the Poisson formula. From this, the probability of obtaining ≥1 events in males if the underlying incidence in the Romero-Gomez study is the same as the pooled incidence is 0.47, therefore adding 0.5 would not seem unreasonable. The equivalent probability for the study by Nesher is 0.51.

In light of the extreme heterogeneity observed in the initial analyses, a further subgroup analysis was performed by study setting (medical centre-based versus population-based). All statistical analyses were performed in R version 4.0.2 (packages *tidyverse, meta, metafor*).

## Results

The process for study selection is shown in a PRISMA flow diagram in Figure 1. The initial database search returned 735 results. After duplicates were removed 519 papers remained, which were then screened. At the title and abstract stage, 505 papers were removed, leaving 14 for the full-text screening.

**Figure 1.**
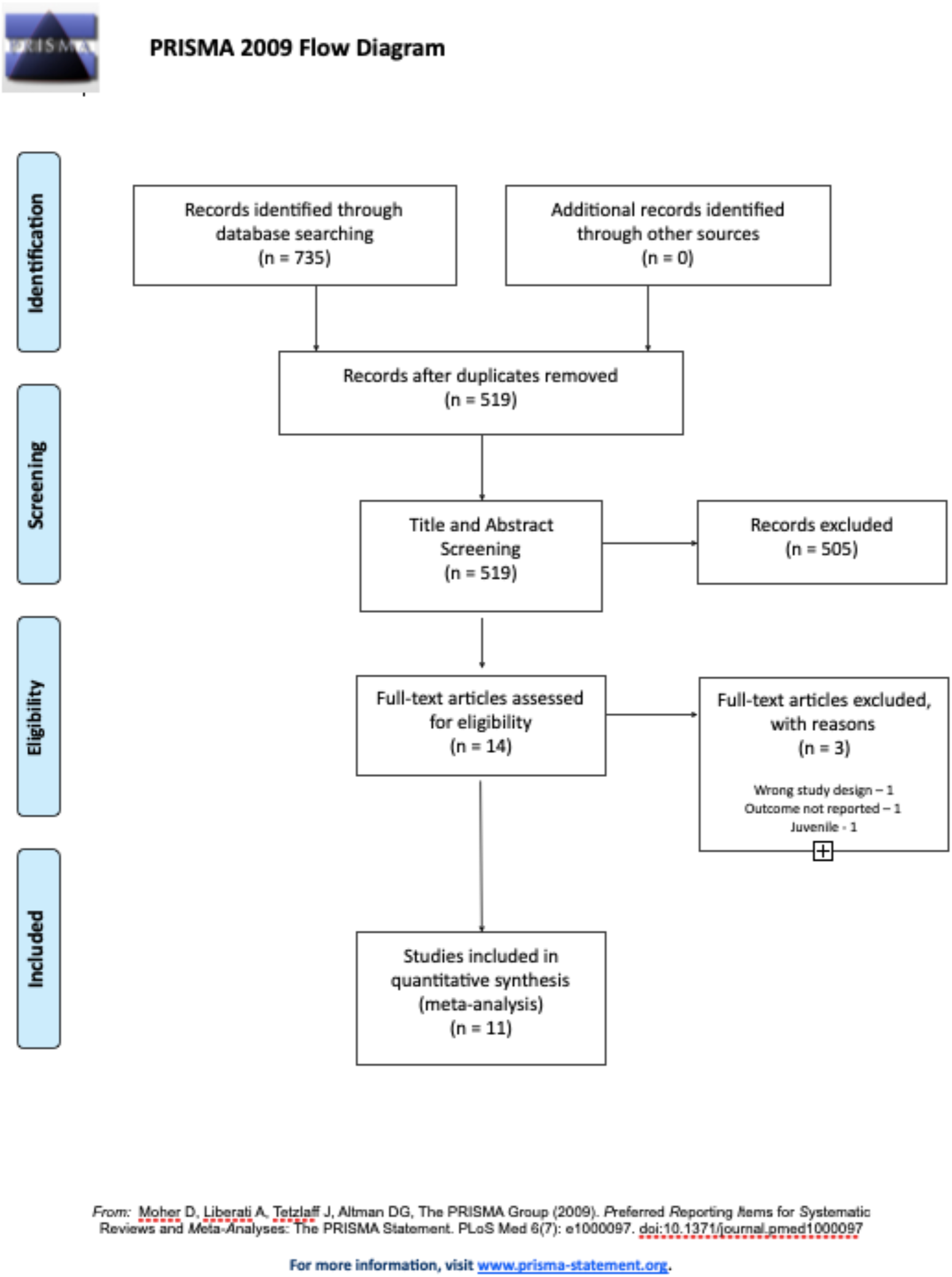
PRISMA flow diagram for study screening and selection for Systematic Review and Meta-analysis of TAK.

Three papers were excluded at the full-text screening stage. One paper was a review and was therefore the wrong study design. One did not report the incidence rate for TAK, nor the background information that would have allowed us to calculate it. Finally, one was excluded as it investigated the incidence of TAK in children only.

Eleven studies met the inclusion criteria for the meta-analysis. Supplementary Table 1 provides an overview of their characteristics.

**Supplementary Table 1.** Summary of the characteristics of studies on incidence of TAK eligible for systematic review and meta-analysis.

| Author(s) | Year | Country | Study Design | Setting | Sample Size | Female: Male | Age at Diagnosis<sup>†</sup> | Quality Score | Diagnostic criteria |
| --- | --- | --- | --- | --- | --- | --- | --- | --- | --- |
| Watts et al | 2009 | UK | Cohort | Population-based | 14 | 13:1 | 51 | 3 | UKGPRD |
| Dreyer, Farschou and Baslund | 2011 | Denmark | Cohort | Population-based | 19 | 16:3 | 36<sup>†</sup> | 3 | Modified Ishikawa |
| Birlik et al | 2015 | Turkey | Cohort | Centre-based | 41 | 38:3 | 37.2 | 2 | ACR |
| Mohammed and Mandl | 2015 | Sweden | Cohort | Centre-based | 13 | 13:0 | 23<sup>†</sup> | 3 | ACR |
| Romero-Gomez et al | 2015 | Spain | Retrospective Cohort | Centre-based | 5 | 5:0 | 26 | 1 | ACR |
| Nesher et al | 2016 | Israel | Retrospective Cohort | Centre-based | 11 | 11:0 | 34 | 2 | ACR or CHCC |
| Saritas et al | 2016 | Turkey | Cohort | Centre-based | 23 | 19:4 | Not reported | 2 | ACR |
| Gudbrandsson et al | 2017 | Norway | Cohort | Population-based | 78 | 68:10 | 33.9 | 3 | ACR or modified Ishikawa |
| Makin, Isbel and Nossent | 2017 | Australia | Retrospective Cohort | Centre-based | 13 | 13:0 | 39<sup>†</sup> | 2 | ACR |
| Park et al | 2017 | South Korea | Cohort | Population-based | 612 | 497:115 | 46 | 3 | RID |
| Kanecki et al | 2018 | Poland | Cohort | Population-based | 177 | 154:23 | 45.4 | 2 | Not performed |

Age: † denotes median, otherwise given as mean. UKGPRD = United Kingdom General Practice Research Database read code for TAK. Modified Ishikawa = Modified Ishikawa diagnostic criteria for TAK. ACR = 1990 American College of Rheumatology diagnostic criteria for TAK. CHCC = Chapel Hill Consensus Criteria for TAK. RID = Rare Intractable Disease registration programme criteria.

Eight of the studies utilised a cohort design, the other three a retrospective cohort design. Five of the studies derived their data from population-based databases, two were single centre based studies and the remaining four included multiple centres. The countries of origin were Turkey, Denmark, Norway, Poland, Australia, Sweden, Israel, South Korea, Spain and the UK. It is notable that most of the data came from Europe (especially Scandinavia) and that only one study (Park et al) looked at the incidence of TAK in Asia, where it was originally described.

Seven of the studies identified patients using local or national databases containing ICD-8, - 9 or −10 diagnostic codes, depending on the time period of the study. Makin et al, 2017, used a mixture of ICD-10 diagnostic codes, key word searches in records of outpatient communication and direct contact with relevant local specialists to identify patients. One study (Watts et al, 2009) used the Read code for TAK used in the national UK General Practice Research Database. One study (Park et al, 2017) identified patients from a national database in Korea called the Rare Intractable Disease (RID) registration programme. Nesher et al (2016) identified patients from local clinical databases.

Confirmation of TAK diagnosis was performed by 10 of the 11 studies, using a variety of methods. Seven used the 1990 American College of Rheumatology (ACR) diagnostic criteria, one the modified Ishikawa criteria and one a combination of the two. One study included patients fulfilling either the 1990 ACR diagnostic criteria or the Chapel Hill Consensus Criteria (CHCC) for TAK. Park et al used the RID diagnostic criteria for TAK, which are similar to the ICD-10 diagnostic criteria other than for age. Kanecki et al (2018) accessed anonymized patient data and did not confirm the diagnosis from patient records.

The meta-analysis of the incidence rate for Takayasu arteritis is shown in the forest plot in Figure 2. The random effects meta-analysis estimated that the pooled-incidence rate of TAK was 1.11 per million person years (95% CI 0.75 – 1.65).

**Figure 2.**
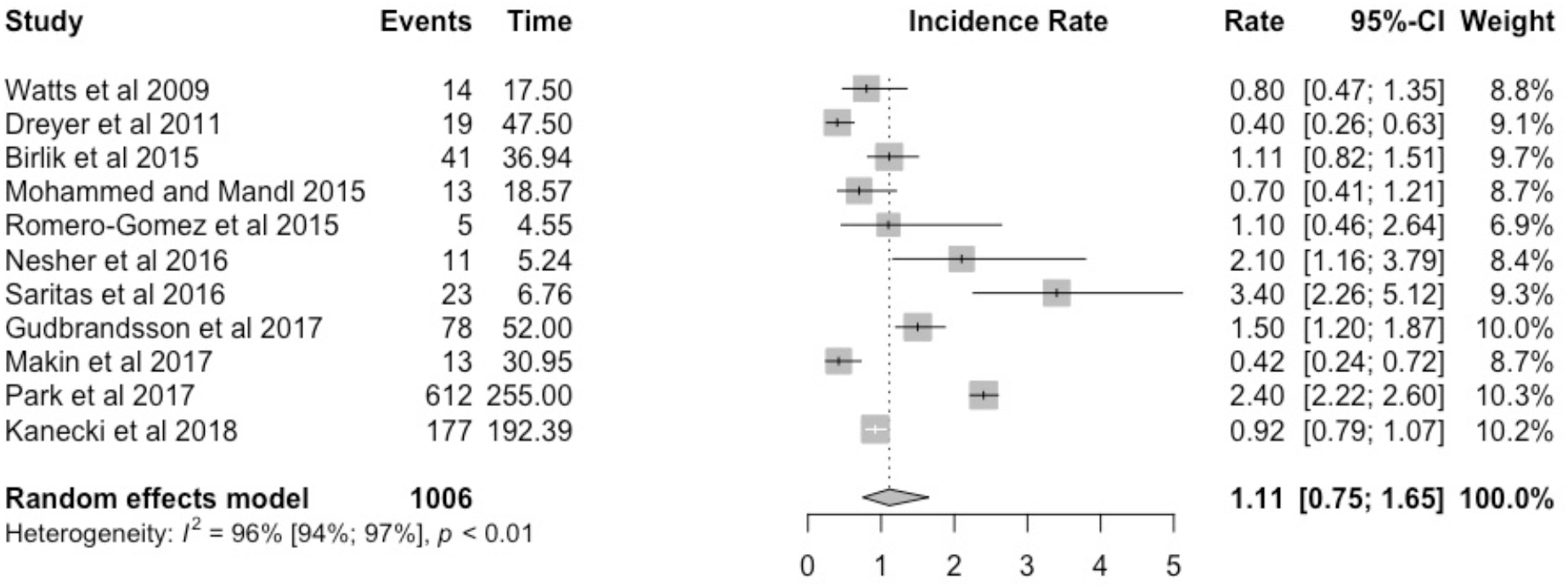
Forest plot showing incidence rate of TAK expressed per million person-years, with 95% confidence intervals.

There was high heterogeneity among the studies pooled for the meta-analysis (I^2^ =96%). The I^2^ statistic describes the percentage of variation across studies that is due to heterogeneity rather than chance.

A subgroup analysis was then performed by sex. The results are shown in a forest plot in Figure 3. The pooled incidence rate was higher for women (2.01 per million person-years, 95% CI 1.39-2.90) than for men (0.28 per million person-years, 95% CI 0.14-0.54). The heterogeneity also remained high with I^2^ values of 94% (women) and 85% (men).

**Figure 3.**
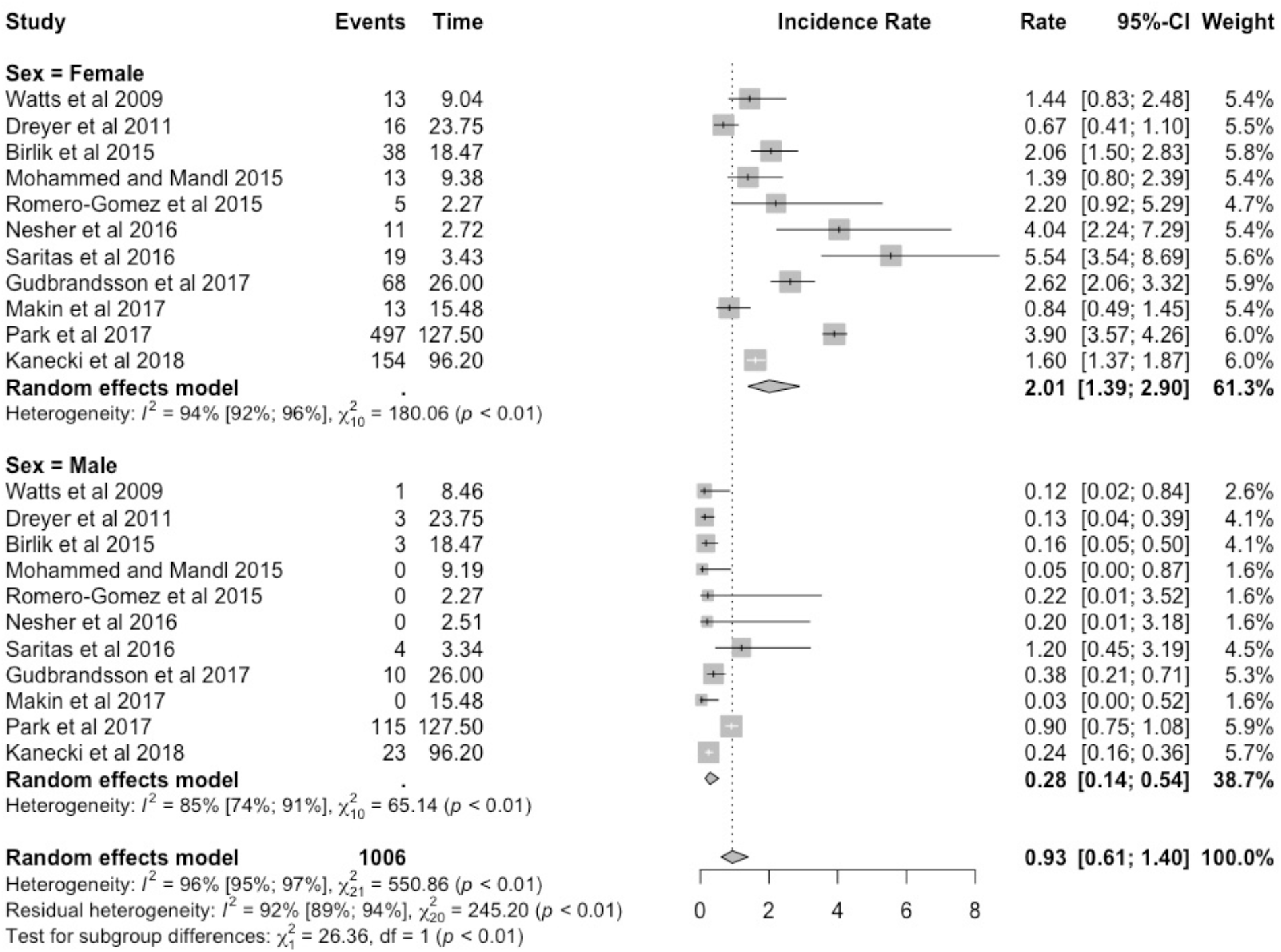
Forest plot of the subgroup analysis by sex, with incidence of TAK expressed per million person-years, with 95% confidence intervals.

The results grouped by quality assessment are presented in a forest plot in Figure 4. Out of a total of 5 possible points for quality, no papers scored 5 or 4, five papers scored 3, five papers scored 2, and one paper scored 1. The incidence rates with 95% confidence intervals were 1.02 (0.61-1.73) and 1.21 (0.69-2.14) respectively for papers that scored 2 and 3. Given that the 95% confidence intervals overlapped, any difference in incidence rate may be due to chance. Heterogeneity remained high with I^2^ values of 92% and 95% n the 2- and 3-point groups respectively.

**Figure 4.**
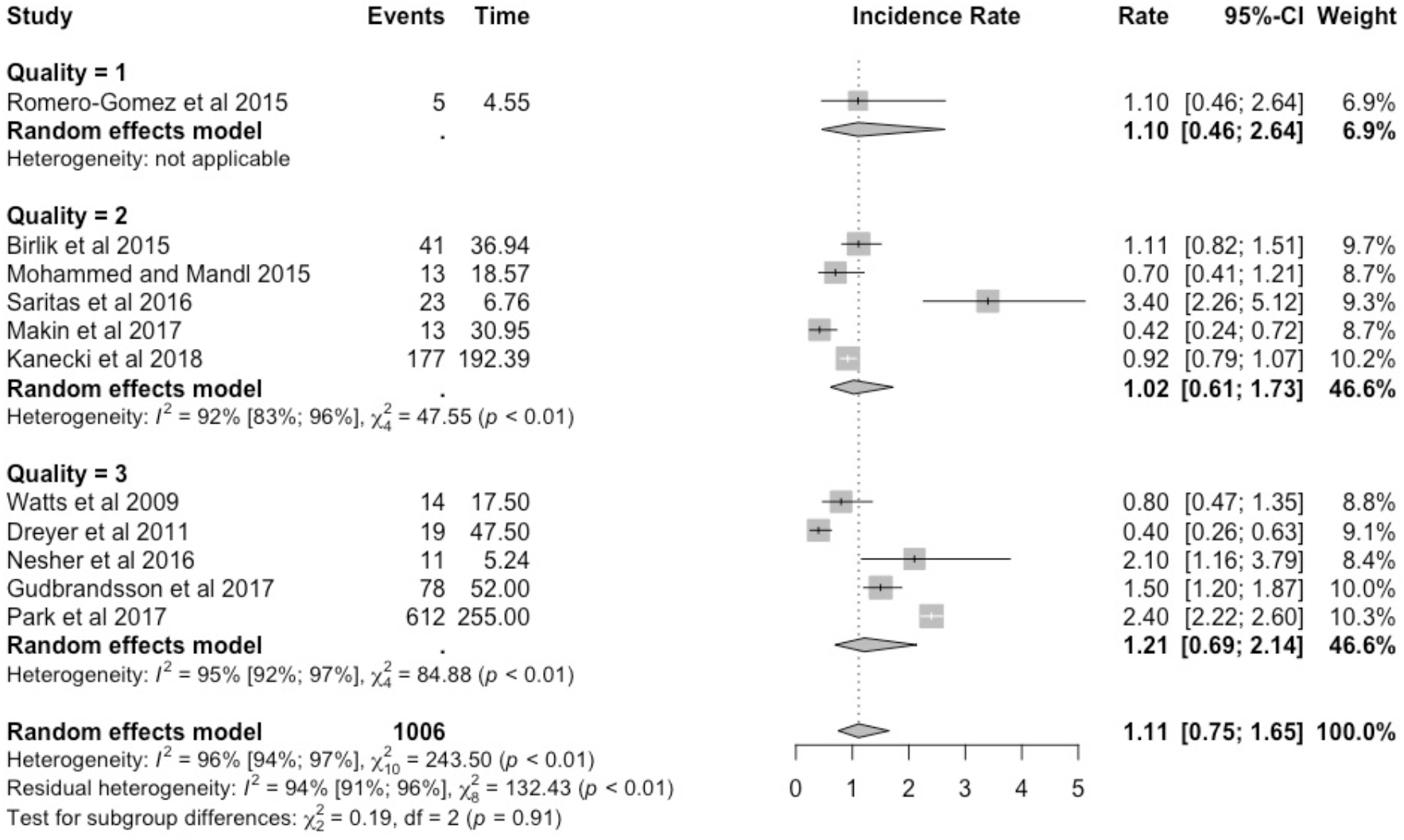
Subgroup analyses based on quality, with incidence of TAK expressed per million person-years, with 95% confidence intervals.

The studies included in this meta-analysis were classified as medical centre-based or population-based settings, and separate subgroup analyses based on these categories are shown in the forest plot in Figure 5. For medical centre-based studies, the incidence rate and corresponding 95% confidence intervals were 1.18 (0.64 – 2.19) and for population-based studies 1.04 (0.58-1.86). The confidence intervals overlap, so the difference in the incidence rates may be due to chance. The high heterogeneity observed remained in both sub-groups, with I^2^ values of 89% for centre-based studies and 98% for population-based studies.

**Figure 5.**
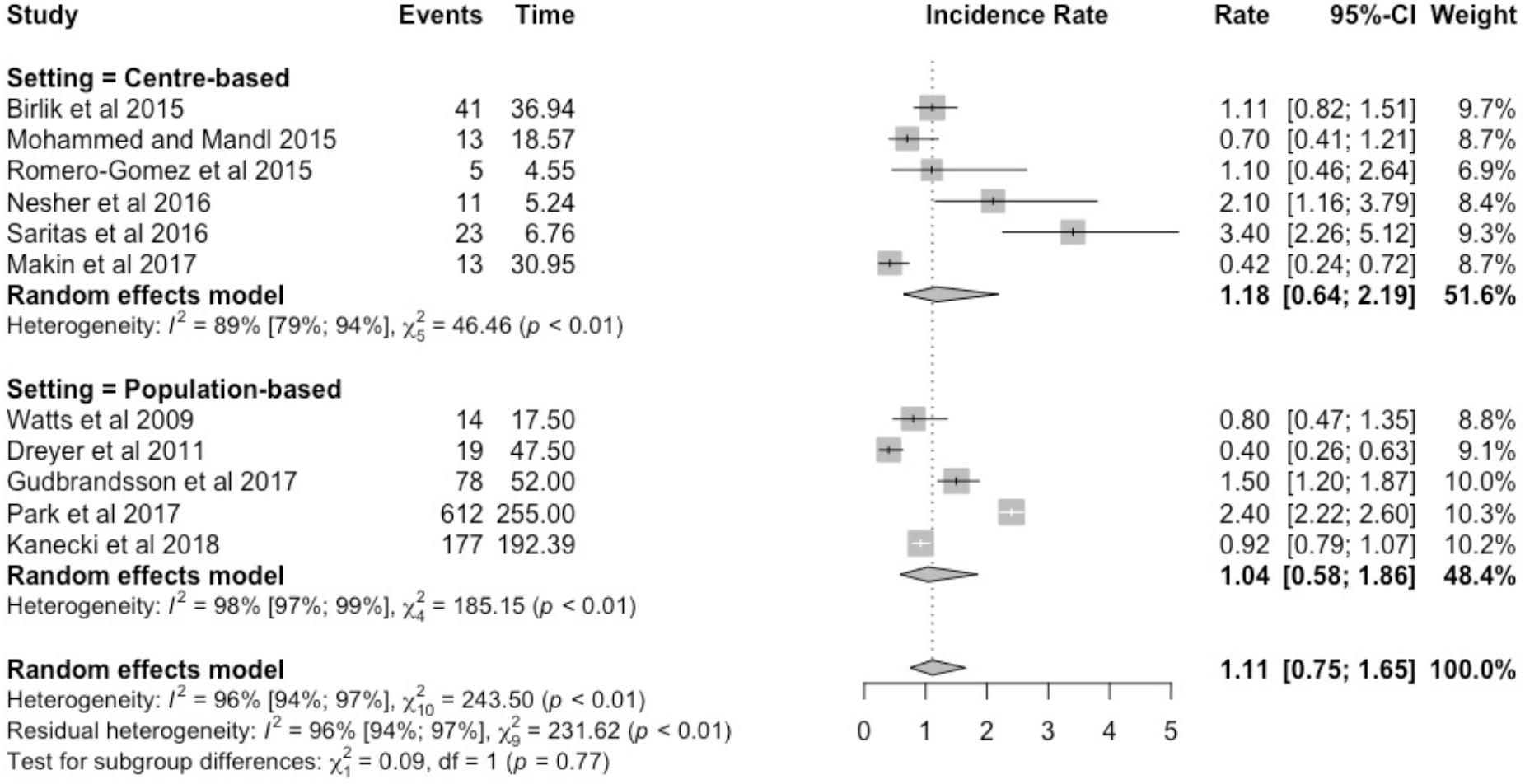
Forest plot of the subgroup analysis based on setting, with incidence of TAK expressed per million person-years, with 95% confidence intervals.

The studies were also analysed according to geographical region and the results are shown in Figure 6. The incidence rate and corresponding 95% confidence intervals were 0.85 (0.59-1.22) for European studies and 1.55 (0.88-2.72) for studies from the rest of the world. The confidence intervals overlap, so the difference in the incidence rates may be due to chance. The high heterogeneity observed remained in both sub-groups, with I^2^ values of 84% for Europe and 94% for the rest of the world.

**Figure 6.**
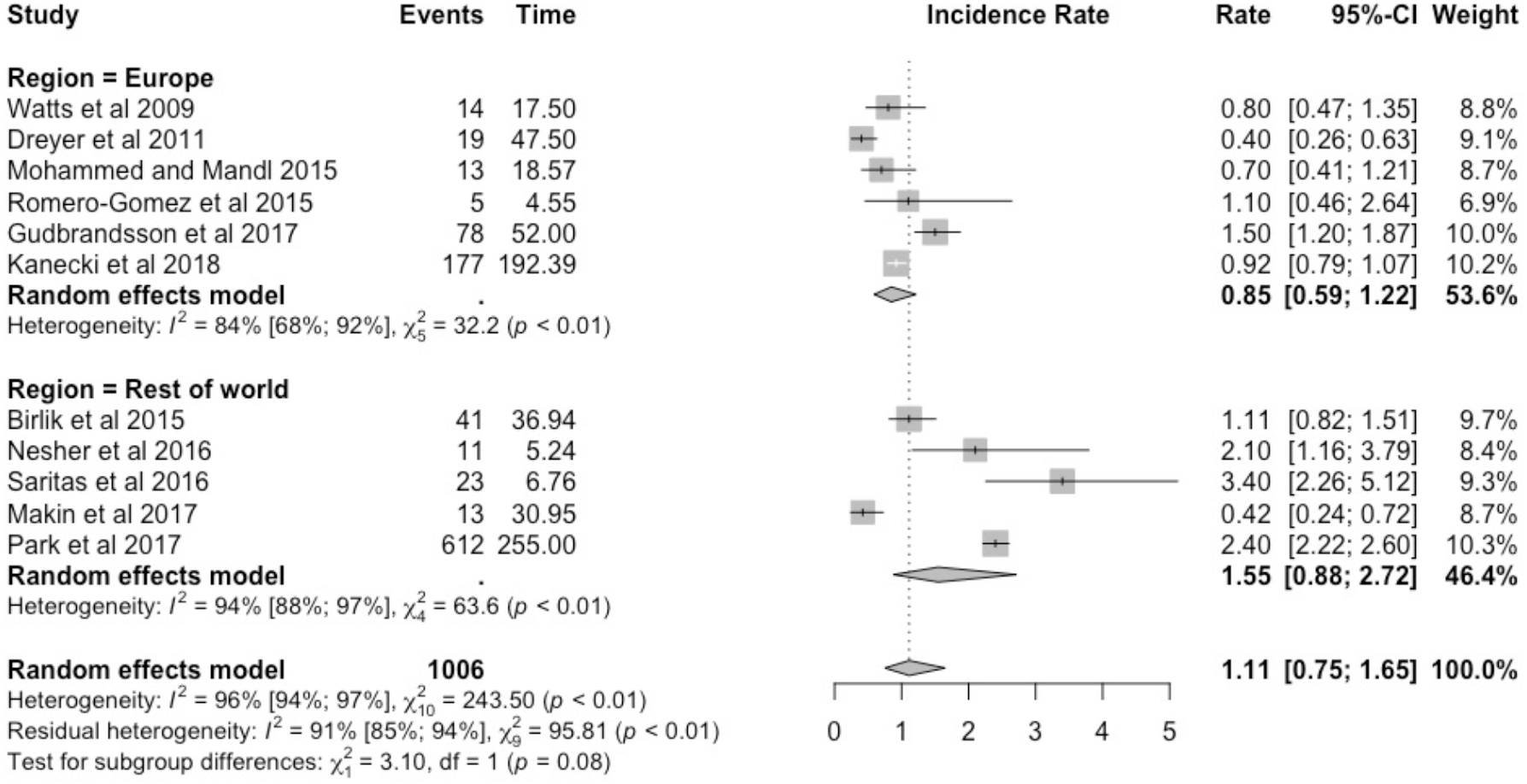
Subgroup analyses of studies based on geographical location, with incidence of TAK expressed per million person-years, with 95% confidence intervals.

Publication bias was assessed using a Begg’s funnel plot which is shown in Figure 7. The results were distributed symmetrically suggesting a lack of publication bias. Seven of the studies fell outside the pseudo 95% confidence intervals.

**Figure 7.**
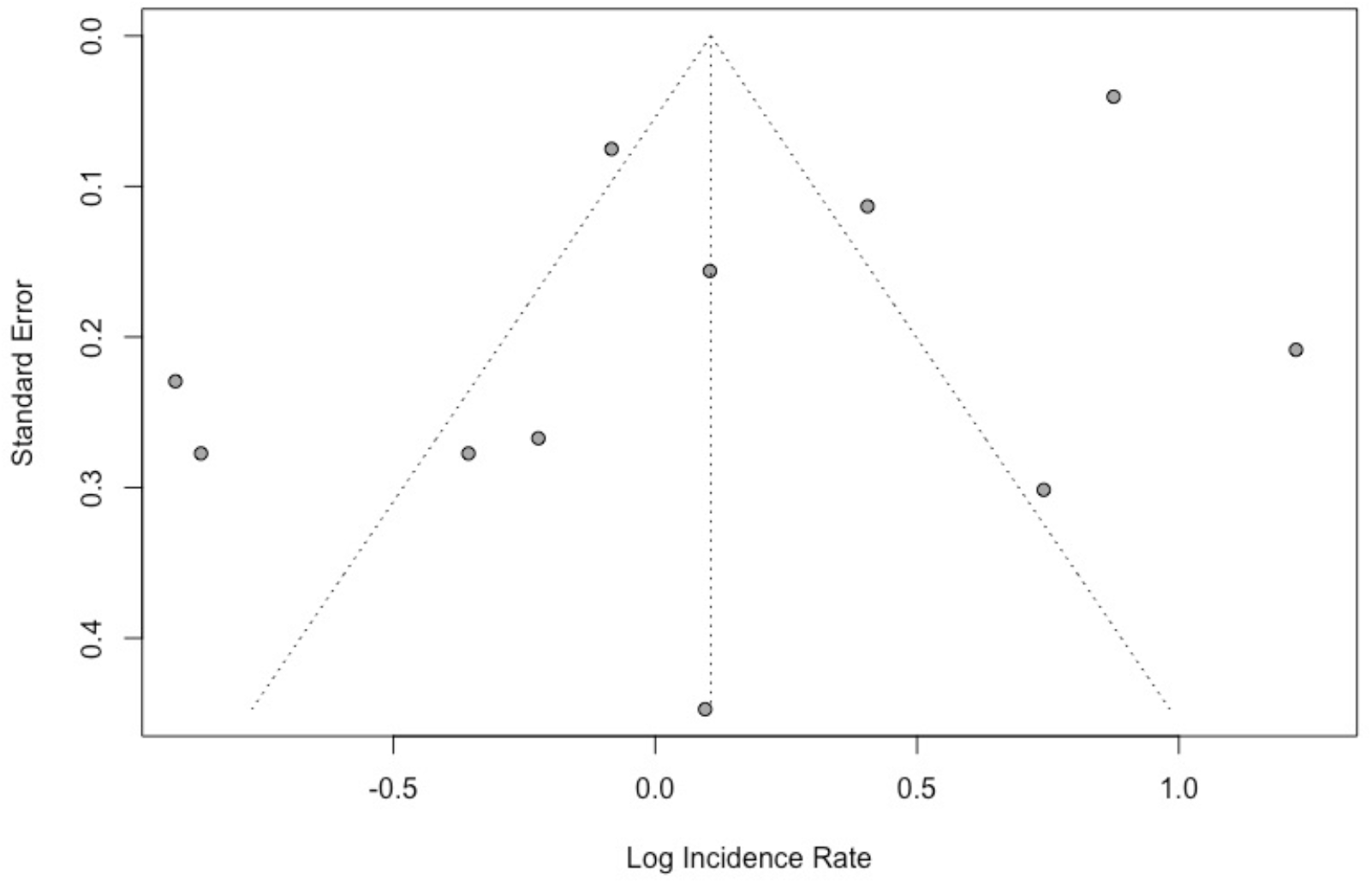
Begg’s funnel plot of the studies investigating the incidence of TAK included in the meta-analysis.

## Discussion

### Main findings

This study is, to the best of our knowledge, the first to examine the global incidence of TAK. The meta-analysis conducted shows TAK to be an exceedingly rare disease with an incidence rate of 1.11 (95% CI 0.75 – 1.65) per million person-years. Our point estimates suggest that TAK is 6 times more common in women than men.

### Heterogeneity in the literature

Whilst TAK is exceedingly rare in all studied populations, there was high heterogeneity in incidence rates between studies. We considered whether this was due to methodology by conducting sub-group analyses by research setting (medical centre- or population-based) and research quality. These analyses did not reduce the heterogeneity seen. The heterogeneity was partially explained by sex but remained high in both the male and female sub-groups. This suggests a true difference in the incidence of TAK between different populations. Data were not available to analyse by race.

### Study design and limitations

To our knowledge, this is the first systematic review of studies of the incidence of TAK. This work adhered to the PRISMA format in the selection and screening of studies used.

Studies were restricted to those published in English where no suitable translation was available. Whilst this is fairly common practice with systematic reviews, particularly due to the increasing shift to publishing in English internationally, it is possible that relevant studies could have been missed. No grey literature was searched which could have provided additional studies for screening and potential inclusion.

Sub-analysis by race was not performed due to a paucity of studies that reported the relevant information. This analysis could have been of benefit as there is evidence that the disease varies in incidence by racial group(3).

This review was limited by the quality of the studies included in the meta-analysis. In our adapted Newcastle-Ottawa scale, the maximum score was 5, which no studies achieved. Most studies scored either 2 or 3, indicating average or poor quality.

The extreme heterogeneity shown in all forest plots was not limited by sex, study quality, setting or geographical region. This demonstrates the limited utility of attempting to calculate global incidence rates for diseases which are likely to vary in their incidence in different populations. We suggest that future epidemiological studies focus on discrete populations and do thorough sub-group analyses to further assess the factors that lead to variation.

### Further research

There is a need for research that determines the incidence of TAK in different geographical areas. At the moment the research is dominated by data from Northern Europe and Turkey. We did not identify any studies from South America or Africa. Sub-analysis of geographical area showed a trend towards lower incidence rates in Europe when compared to the rest of the world. However, the overlap in confidence intervals means that this could have occurred through chance.

### Clinical implications

Tocilizumab has recently been commissioned by NHS England as a third line treatment for TAK in adults “where attempts to control disease progression of TAK have failed”(5). It is estimated that up to 50% of patients with TAK may benefit from a biologic agent(5). The commissioning policy is based on the incidence of TAK in Southern Sweden, from 13 patients identified between 1997 and 2011 in a population of almost 1 million(8). In this way, TAK is an exemplar for many rare rheumatic diseases where we lack epidemiological data to guide healthcare planning for conditions that require highly specialised treatment and concentrated clinical knowledge and resources.

To provide more accurate incidence data for these rare diseases, we need contemporary whole population-based studies. This has recently been made possible by the creation of the National Congenital Anomaly and Rare Disease Registration Service by Public Health England and the Registration in Complex Rare Diseases – Exemplars in Rheumatology (RECORDER) project, which aims to identify and build a register of those individuals living with rare diseases, including rare autoimmune rheumatic diseases, amongst the 55 million people living in England(9, 10).

## Conclusions

This systematic review of the literature has demonstrated that the incidence rate of TAK is 1.11 (95% CI 0.75-1.65) cases per million-person years. TAK is more common in women than men. There appears to be a genuine difference in incidence in different populations. The reasons for this warrant further research.

## Data Availability

Data are available upon reasonable request

## Key Messages

1. Takayasu arteritis (TAK) is an ultra-rare disease, which has implications on diagnostic pathways, service design, funding of treatment and clinical trials.
2. TAK is more common in women than men
3. Incidence of TAK differs across different populations
4. This research highlights the value of accurate incidence data and whole population epidemiology when designing and commissioning clinical services and treatments

## Appendix 1. Adapted Newcastle-Ottawa scale

### COHORT STUDIES

<u>Note</u>: A study can be awarded a maximum of one star for each numbered item.

#### Selection

1. Representativeness of the cohort
  a. truly representative ✵
  b. somewhat representative ✵
  c. no description of the derivation of the cohort
2. Ascertainment of exposure
  a. secure record (e.g. surgical records) ✵
  b. written self-report
  c. no description
3. Demonstration that outcome of interest was not present at start of study
  a. yes ✵
  b. no
4. Was the study population large enough?
  a. >1,000,000 ✵
  b. <1,000,000

#### Comparability

1. Comparability of cohorts on the basis of the design or analysis
  a. Study controls for gender, ethnicity or age (select the most important factor) ✵

## Appendix 2. Data extraction form

|  |  |  |  |  |  |  |  |  |
| --- | --- | --- | --- | --- | --- | --- | --- | --- |
| Author(s) | Year | Type of Study | Setting | Population | Age | incidence/1.000,000 | LCI | UCI |
| Female:male | logir | loguci | loglci | Quality | Male ir | Male uci | Male lci | Male logir |
| Male loguci | Male loglci | Female ir | Female uci | Female lci | Female logir | Female loguci | Female loglci |  |

